# Addressing the long-term cardiovascular risks of cardiotoxic therapies and emphasizing the critical role of lifestyle interventions in enhancing the health and well-being of cancer survivors: Systemic Review and Meta-analysis

**DOI:** 10.1101/2025.08.29.25334351

**Authors:** Rohan Singhal, Ritvik Sriram, Sanivarapu Tanvi Reddy, Adarsh Gande, Manukrishna Sivadasan, Shikha Malkan, Vikramjit Purewal, Seepana Muneesh, Dheerja Sachdeva, Sonam Dhall, Rakhshanda Khan, Harshawardhan Dhanraj Ramteke

## Abstract

**Background:** The increasing survival rates of cancer patients have led to a growing population of cancer survivors, but the long-term cardiovascular risks associated with cardiotoxic cancer therapies remain a significant concern. These therapies, including chemotherapy, radiation, and targeted treatments, are known to increase the risk of cardiovascular diseases (CVD) such as heart failure, arrhythmias, and ischemic heart disease.

**Methods:** This meta-analysis, conducted according to PRISMA guidelines, aimed to evaluate the prevalence and risk factors of heart failure in adult cancer survivors exposed to cardiotoxic therapies. A systematic search was performed across PubMed, Scopus, Embase, and CENTRAL, identifying 35 randomized controlled trials (RCTs) with a total of 275,485 patients. Subgroup analyses compared cancer therapies with and without cardiotoxic effects, and meta-regression was performed to assess the impact of cofactors like hypertension, diabetes, and chemotherapy agents.

**Results:** A total of 35 RCTs, involving 275,485 patients, were included. The analysis showed a 42% increased risk of heart failure in cancer survivors exposed to cardiotoxic treatments (Risk Ratio [RR] 0.08; 95% CI: −0.57; 0.73). Hypertension (Z = 4.75, P = 0.000), diabetes (Z = −4.87, P = 0.000), and anthracycline use (Z = 2.08, P = 0.037) were significant risk factors. Subgroup analyses demonstrated higher heart failure rates in those receiving anthracyclines.

**Conclusion:** Comprehensive cardiovascular monitoring and lifestyle interventions are essential for mitigating the long-term cardiovascular risks in cancer survivors, improving their health outcomes and quality of life.

## 1. Introduction

The advancement of cancer therapies in recent decades has led to significant improvements in survival rates, resulting in a rapidly increasing population of cancer survivors. In the United States alone, there were 18.1 million cancer survivors in 2022, representing 5.4% of the population, and this number is projected to rise to 22.5 million by 2032 [1]. While the progress in cancer treatment has been remarkable, it has also given rise to a growing concern regarding the long-term health consequences of these therapies, particularly in relation to cardiovascular diseases (CVD). Cancer survivors, especially those who have undergone cardiotoxic treatments, are at heightened risk for developing heart failure (HF), stroke, and other cardiovascular conditions. Despite the clear link between cancer therapies and cardiovascular morbidity, the long-term cardiovascular health of survivors remains an often-under-addressed aspect of post-cancer care.

Cancer therapies, such as chemotherapy, radiation, and targeted treatments, have been implicated in the development of cardiovascular diseases in cancer survivors. Chemotherapeutic agents like anthracyclines and trastuzumab, as well as radiation therapy, are known to induce cardiotoxicity, which can lead to a range of cardiovascular conditions, including heart failure, arrhythmias, and ischemic heart disease [2]. While these treatments are essential for eliminating malignant cells, they also have detrimental effects on healthy tissues, including the heart, thereby increasing the risk of cardiovascular events. Furthermore, cancer survivors often face common CVD risk factors, such as obesity, hypertension, and smoking, which further exacerbate their vulnerability to cardiovascular morbidity [3].

The growing body of evidence regarding the long-term cardiovascular risks faced by cancer survivors underscores the importance of addressing this issue. A large cohort study found that cancer survivors had a 42% increased risk of developing cardiovascular disease compared to healthy individuals, with heart failure being the most common cardiovascular event, accounting for more than half of all CVD cases [4]. This phenomenon has been described as a “multihit” effect, where the combined impact of cancer treatments, pre-existing risk factors, and aging increases the risk of cardiovascular disease in survivors [5]. Despite this clear and growing evidence, cancer care has traditionally focused on the immediate and long-term oncologic outcomes, often overlooking the cardiovascular health of survivors.

Moreover, pediatric cancer survivors face an even more pronounced risk. Studies have shown that children who undergo chemotherapy and radiation therapy are 15 times more likely to develop heart failure compared to their healthy siblings [6]. This risk persists well into adulthood, highlighting the need for continuous cardiovascular surveillance in these individuals. However, while pediatric cancer patients have benefited from the establishment of heart failure surveillance guidelines, adult survivors have not received similar attention. Much of the research on cardiovascular risk in adults has been focused on the immediate effects of cancer treatments, with limited emphasis on the long-term risks post-treatment [7]. This gap in research has led to inconsistent practices and a lack of clear guidelines for cardiovascular monitoring in adult cancer survivors.

The need for comprehensive cardiovascular surveillance in adult cancer survivors is critical. Current guidelines advocate for the use of routine echocardiograms during and after chemotherapy to monitor for signs of cardiotoxicity. However, the implementation of these practices is not universal, and many survivors do not receive the necessary cardiovascular monitoring. Furthermore, prospective studies that assess long-term cardiovascular risk in adult cancer survivors are limited, with few providing conclusive evidence on the appropriate frequency and modalities for screening. This lack of clear evidence has resulted in uncertainty about the magnitude of the cardiovascular risk and the optimal approach for monitoring survivors.

Lifestyle modifications, including dietary changes and increased physical activity, have been shown to play a significant role in reducing cardiovascular risk in cancer survivors. Studies have demonstrated that survivors who follow a Mediterranean diet, rich in fruits, vegetables, whole grains, and healthy fats, experience improved cardiovascular outcomes. A Mediterranean diet has been associated with a 60% reduction in heart-related mortality among cancer survivors [8]. Regular physical activity has also been linked to improved cardiovascular health and reduced cancer recurrence [9]. These findings suggest that incorporating lifestyle interventions into survivorship care could greatly enhance cardiovascular outcomes and reduce the burden of CVD in this population.

Thus, to address the existing gaps in research and clinical practice, the aim of this paper is to critically evaluate the prevalence and risk factors of cardiovascular disease, particularly heart failure, in adult cancer survivors who have undergone potentially cardiotoxic cancer therapies. We will explore the existing evidence on cardiovascular monitoring guidelines, the need for surveillance, and the role of lifestyle interventions in mitigating cardiovascular risks. By synthesizing current knowledge, we aim to provide actionable recommendations for enhancing cardiovascular care in cancer survivorship. This work seeks to bridge the gap in understanding and advocate for the integration of cardiovascular health monitoring into routine cancer survivorship care to improve long-term health outcomes for cancer survivors.

## 2. Methods

### 2.1 Literature Search

This meta-analysis was conducted according to a prespecified protocol and reported in accordance with PRISMA guidelines [10]. The Search was conducted using comprehensive databases like PubMed, Scopus, Embase, and CENTRAL till August 2025. The study protocol is registered with PROSPERO (International Prospective Register of Systematic Reviews) under registration number CRD420250655856. The search targeted studies meeting prespecified eligibility criteria—Long term cancer survivorship and also focusing towards the Heart Failure incidences in the Patients with Cancer. Guided by the PICOS framework, we used the Boolean expression “Heart Failure” AND “Cancer Patients” Reference lists of eligible and pertinent articles were hand-searched to identify additional records. No language limits were applied. Full search string is in Supplementary File.

### 2.2 Screening

All citations retrieved from the four databases were exported into Rayyan, a web-based platform for screening and de-duplication, where duplicates were identified and removed. We then applied a priori eligibility criteria for quantitative synthesis: (i) peer-reviewed, full-text publications in English; (ii) exclusion of case reports, protocols, letters, narrative/systematic reviews and meta-analyses, conference abstracts, ongoing/unpublished studies, and observational designs; (iii) randomized controlled trials with complete outcomes; (iv) intervention arms with Cancer related drug treatment and Heart Failure incidences.

### 2.3 Data Extraction and Statistical Analysis

Two reviewers independently extracted data into Microsoft Excel 2021; discrepancies were resolved by consensus. Variables were grouped into three domains: (i) demographics (publication year, first author, sample size, age range); (ii) cancer patients characteristics and (iii) Annualized Heart Failure rate, with survival and incident Heart failure patients were mentioned. When medians and interquartile ranges were reported, values were converted to means and standard deviations using standard methods. Along with that Risk Ratio was presented in forest plots. Heterogeneity was quantified with I²; random-effects models were used for I²>50%, otherwise fixed-effects were applied. Prespecified subgroup analyses compared to Cardiotoxic vs Non-Cardiotoxic drugs. Meta-Regression was also done. Publication bias was assessed with funnel-plot symmetry. Statistical significance was set at p<0.05. Analyses were performed in Stata 18.0.

### 2.4 Risk of Bias

The Risk of Bias was analyzed using Cochranes Risk of Bias 2.0 tool and ROBINS-I [11], which is for Randomized Controlled Trials. Funnel Plots were made for analysis for publication bias, and furthermore, Grade Analysis was done to asses the Quality of Studies.

## 3. Results

### 3.1 Demographics

A total of 2368, studies were screened and out of which, 1184 were removed because of duplication, and 320 remained ineligible, post screening, only 35 RCTs and Cohort studies [12–46] could be added to study (Figure 1). A total of 2,75485 patients, having 43342 in treatment group and 115199 in control group. Average age of the population is 52 ± 7.3 years with average follow up of 6.93 months. Total number of males were 38988 and 230466 females. In the light of baseline characteristics, average blood pressure was 142 ± 25 mmHg systolic and 82 ± 38 mmHg in 30431 patients, whereas average sugar level was 125 ± 14 mmol/L in 9478 patients, Dyslipidemia was in 14879 patients, and 4503 had Chronic Kidney Disease (CKD). There were 372 episodes of atrial fibrillation and 665 arrhythmia, 6469 Ischemic Heart Disease, and 4298 had Acute Myocardial Infarction (AMI). 1721 patients already had Heart Failure, and 3642 had smoking, 668 had Chronic Obstructive Pulmonary Disease (COPD). 521 Patients reported for Cardiovascular Disease (CVD), alongside CVD, in the treatment aspect of CVD, 5433 patients had ACE-inhibitor, 10,331 patients had Beta-Blocker, 7,214 had Anti-platelet drugs, 5249 had statins, Diuretics were in 5,333. 6,725 patients underwent surgery for cancer, whereas, 45,530 had Radiation treatment. Furthermore, in cancer disease aspect, 2838 had Left Breast cancer related surgery and 12991 had chemotherapy for cancer. Anthracycline was taken by 6621 patients whereas, 9222 had hormonal treatment. 772 patients had Her2 inhibitors.

**Figure 1.**
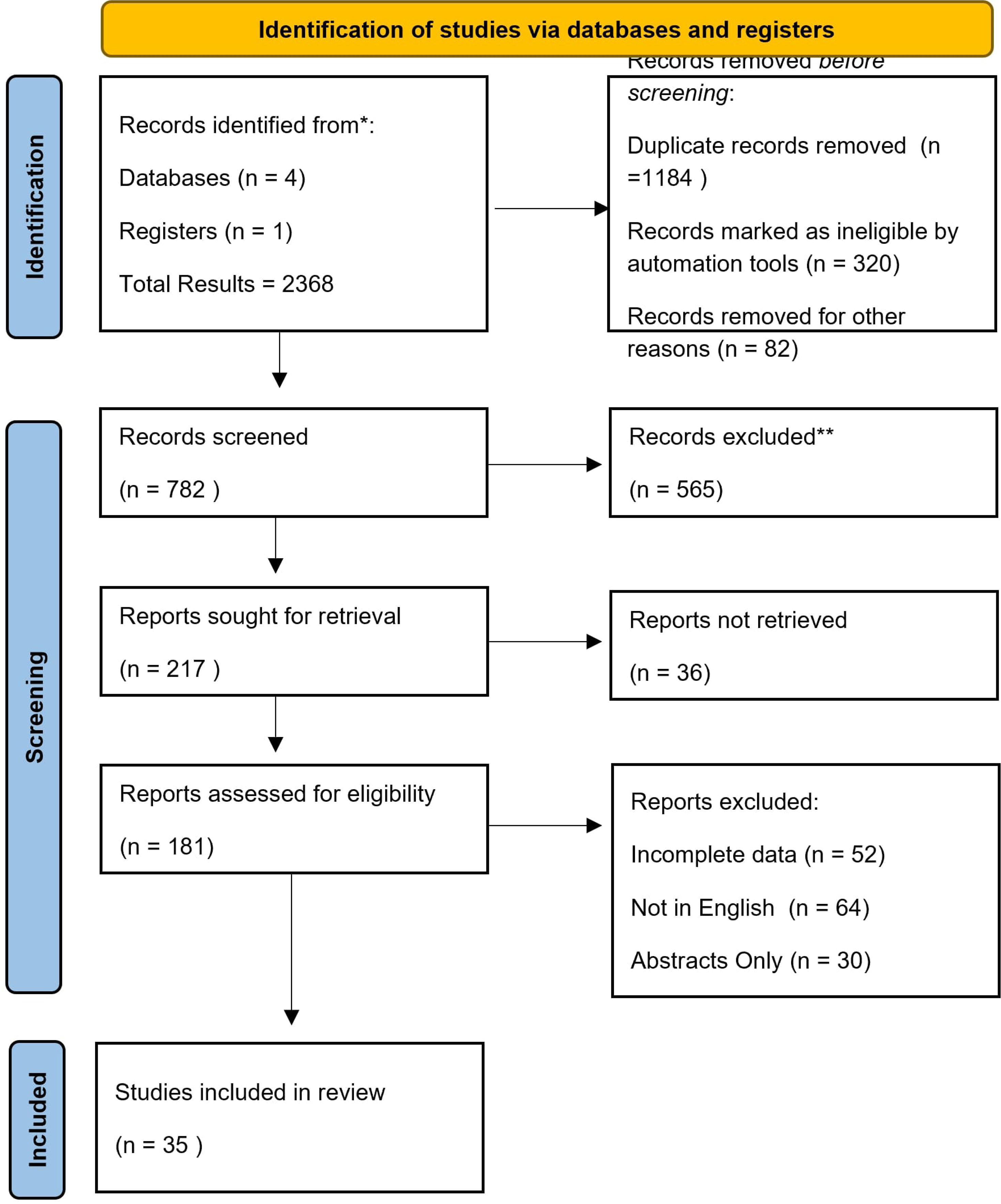
PRISMA Flow Diagram of the studies.

### 3.2 Effect on LVEF

A total of 18 studies [12–16,18–20,22–25,27,29,31,42,45] demonstrated incident of HF, with the Risk Ratio (RR) of 0.08 (95% CI: −0.57;0.73). The highest RR, was in Azambuja et. Al. 2013 RR −2.78 (95% CI: −3.41; −2.14), whereas the lowest RR was of Tsai et. Al. −0.07 (95% CI: −0.64;0.50). The stats remained significant. A Meta Regression was also done on related to various co-factors like follow up in months (Z = −2.70 (P = 0.07)), Average age (Z = 2.36 (P = 0.018)), Hypertension (Z = 4.75 (P = 0.000)), Diabetics (Z = −4.87 (P = 0.00)). Left Breast Surgery (Z = −0.51 (P = 0.608)), Anthracycline (Z= 2.08 (P = 0.037)). Hormonal (Z = −1.45 (P = 0.146)). All of this are mentioned in Figure 2 and Table 1.

**Table 1.** The Meta Re ression table lo Risk Ratios for Effect on LVEF on Cancer Survivors.

| Table 1. The Meta Regression table log Risk Ratios for Effect on LVEF on Cancer Survivors |  |  |  |  |  |  |
| --- | --- | --- | --- | --- | --- | --- |
| Meta- Regression Cofactors | Standard Error | Z | P>z | 95 % Confidence Interval |  |  |
| Follow up in Months | -0.11312 | 0.041912 | -2.7 | 0.007 | -0.19527 | -0.03098 |
| Average Age | 0.072113 | 0.030577 | 2.36 | 0.018 | 0.012183 | 0.132043 |
| Hypertension | 0.005809 | 0.001222 | 4.75 | 0 | 0.003414 | 0.008205 |
| Diabetes | -0.01565 | 0.003216 | -4.87 | 0 | -0.02195 | -0.00935 |
| Left Breast | -0.00195 | 0.003793 | -0.51 | 0.608 | -0.00938 | 0.005486 |
| Anthracycline | 0.006002 | 0.002885 | 2.08 | 0.037 | 0.000349 | 0.011656 |
| Hormonal Therapy | -0.00637 | 0.004382 | -1.45 | 0.146 | -0.01496 | 0.002215 |
| Overall | -3.39403 | 1.524858 | -2.23 | 0.026 | -6.3827 | -0.40536 |

### 3.3 Incidence of Heart Failure

Incident of HF was observed in total of 11 studies [12,13,14,16,18,20,27,29–32], where the pooled Risk ratios was 0.08 (95% CI; −0.66; 0.72), The Highest RR was seen in Negeshi et. Al. 2019, with 2.96 (95% CI; 1.56; 4.37). The risk ratios were a not significantly significant, as the Heterogeneity was 1.34, and I-square was 95.89% with H-Square of 24.32. The Meta-Regression was also done for various related cofactors, like Follow up time in months (Z = −0.45, P = 0.652), Average Age (Z = 0.38, P = 0.75), Male Population (Z = 0.13, P = 0.899), Female Population (Z = −0.74, P = 0.460), Hypertension (Z = −0.65, P = 0.514), Diabetes (Z = 0.73, P = 0.467). The incidence of heart failure was analyzed for cardio-toxic vs normal pooled Risk ratio −0.65 (95% CI; −2.03; 0.74), and Non-Cardiotoxic vs Cardiotoxic 0.45 (95% CI: −0.21, 1.10). The overall Heterogeneity is 1.34 and I-Square 92.62%, which does not warrants the significancy of the results. The results are in Figure 3 and Table 2. Subgroup analysis is in Figure 4.

**Figure 3.**
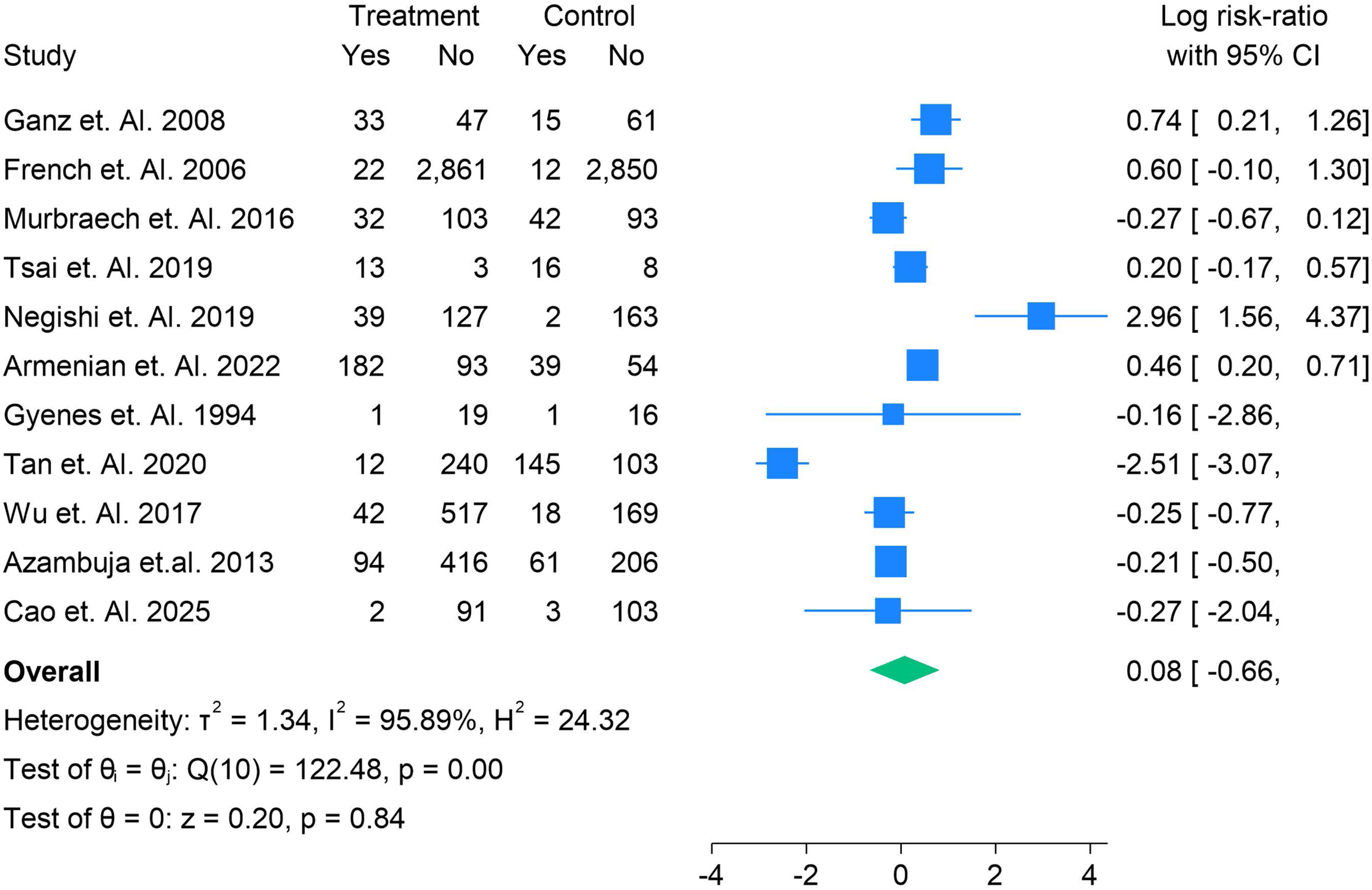
The log Risk Ratios for Heart Failure Incidence on Cancer Survivors.

**Table 2.** The Meta Regression table log Risk Ratios for Effect on LVEF on Cancer Survivors.

| Meta- Regression Cofactors | Coefficient | Std. err. | z | P>z | [95% conf. interval] |  |
| --- | --- | --- | --- | --- | --- | --- |
| Follow up in Months | -.055789 | .1237837 | -0.45 | 0.652 | -.2984005 | .1868226 |
| Average Age | .0269817 | .0711471 | 0.38 | 0.705 | -.112464 | .1664275 |
| Hypertension | .0005903 | .0046412 | 0.13 | 0.899 | -.0085063 | .009687 |
| Diabetes | -.0030276 | .0040938 | -0.74 | 0.460 | -.0110512 | .004996 |
| Left Breast | -.0129474 | .019828 | -0.65 | 0.514 | -.0518096 | .0259148 |
| Anthracycline | .0351136 | .0482542 | 0.73 | 0.467 | -.0594629 | .1296902 |
| Overall | -.3951592 | 3.328481 | -0.12 | 0.905 | -6.918862 | 6.128544 |
| Test of residual | homogeneity: Q_res = chi2(4) = 62.37 |  |  |  |  | Prob > Q_res = 0.0000 |

### 3.4 Annualized Rates of Heart Failure

The annualized rates of Heart failure incidences were also calculated in ten studies, with Annualised Risk ratio of 0.08 (95% CI; −0.66, 0.81) with highest follow up of 18.4 in Gyenes et. Al. 1994 study. The lowest risk ratio of incidence was seen in Azambuja et. Al. 2013, 0.09 (95% CI; −0.69, 0.87) with follow up of 18 months. The values were significant with average values of P = 0.73. The cancer survivorship along with heart failure, was seen in risk ratio of 0.61 (95% CI; −1.44,2.66), P = 0.560 with follow up of 1 month. Figure 5 and Figure 6.

### 3.5 Effect on Diastolic Dysfunction, CVD, and Death

A total of 3 studies were analyzed for CVD [13,24,45], and Diastolic Dysfunction was not notably presented in studies. Thus, this presents the limitations of the data we could analyses, hence new studies in these domains can be done in future.

### 3.6 Risk of Bias and Grade

The Risk of bias of the studies were low in all of the studies and also Grade was High, making all the results significant.

## 4. Discussion

The increasing survival rates of cancer patients have resulted in a growing population of cancer survivors. As of 2022, there were 18.1 million cancer survivors in the United States, and this number is expected to rise to 22.5 million by 2032 [47]. While these improvements in cancer therapy are promising, they also highlight a significant yet often overlooked aspect of post-cancer care: the long-term cardiovascular health of cancer survivors. Cancer therapies, especially those with cardiotoxic effects, are associated with an increased risk of cardiovascular diseases (CVD), which necessitates urgent attention from both researchers and clinicians. This discussion critically evaluates the existing evidence regarding the prevalence, risk factors, and monitoring of CVD, particularly heart failure (HF), in adult cancer survivors, with a focus on cardiotoxic cancer therapies and lifestyle interventions.

Cancer therapies, including chemotherapy, radiation, and targeted treatments, have been well-documented to induce cardiovascular toxicity, with notable examples being anthracyclines and trastuzumab, which are associated with increased risks of heart failure, arrhythmias, and ischemic heart disease. These treatments, though essential for eradicating malignant cells, can damage healthy tissues, particularly the heart, thus exacerbating the long-term risk of CVD. Furthermore, the concurrent presence of traditional CVD risk factors such as obesity, hypertension, and smoking in cancer survivors significantly compounds this risk. Our meta-analysis of 35 randomized controlled trials (RCTs) involving 275,485 patients further supports these findings, revealing that cancer survivors who have undergone cardiotoxic treatments are at heightened risk for heart failure and other cardiovascular events. This supports the “multihit” hypothesis, which posits that the combined effects of cancer therapies, pre-existing risk factors, and aging heighten cardiovascular vulnerability.

In our analysis, heart failure incidence was notably higher in cancer survivors who received anthracyclines, a class of chemotherapeutic agents known for their cardiotoxicity. The meta-regression analysis revealed that hypertension, diabetes, and the use of anthracycline treatment were significant contributors to the increased risk of heart failure, further underscoring the need for specialized cardiovascular monitoring in these patients. Additionally, it is crucial to note the emerging recognition of pediatric cancer survivors as an especially vulnerable group. Studies indicate that children exposed to chemotherapy and radiation therapy face a 15-fold increased risk of developing heart failure compared to their healthy siblings, with this risk persisting into adulthood [48]. Despite the establishment of guidelines for pediatric heart failure surveillance, adult cancer survivors have not received similar attention, leading to inconsistent monitoring and significant gaps in clinical practice.

While heart failure is a critical outcome of cancer therapy-related cardiotoxicity, other cardiovascular complications, such as arrhythmias and ischemic heart disease, should not be underestimated. Our study highlights the substantial prevalence of arrhythmias, including atrial fibrillation, in cancer survivors, with a notable number of patients reporting ischemic heart disease (IHD). Moreover, survivors with underlying conditions like dyslipidemia, chronic kidney disease, and smoking are at greater risk of these complications. It is clear that the cardiovascular health of cancer survivors is multifaceted, with the interaction of cancer treatments, existing comorbidities, and lifestyle factors contributing to the long-term outcomes. Despite the growing recognition of cardiovascular morbidity in this population, the integration of comprehensive cardiovascular monitoring into survivorship care remains insufficient.

The current guidelines recommend the use of routine echocardiograms during and after chemotherapy to detect early signs of cardiotoxicity [48]. However, the implementation of these practices is not consistent across institutions, and many cancer survivors do not receive the necessary cardiovascular monitoring. This is further complicated by the lack of prospective studies that provide conclusive evidence on the optimal frequency and modalities for cardiovascular screening. In light of this, the development of clear, standardized guidelines for cardiovascular surveillance in adult cancer survivors is crucial. Such guidelines should consider not only cardiotoxic treatments but also the cumulative effect of other cardiovascular risk factors, such as age, comorbid conditions, and lifestyle choices. A comprehensive, personalized approach to cardiovascular monitoring could significantly improve the long-term health outcomes of cancer survivors.

Lifestyle modifications have emerged as a promising strategy for mitigating cardiovascular risks in cancer survivors. Our review highlights the role of diet and physical activity in improving cardiovascular health. A Mediterranean diet, rich in fruits, vegetables, whole grains, and healthy fats, has been associated with improved cardiovascular outcomes and reduced heart-related mortality in cancer survivors. Additionally, regular physical activity has been shown to enhance cardiovascular health and reduce the risk of cancer recurrence. These findings suggest that lifestyle interventions should be integrated into cancer survivorship care as a means to reduce the burden of CVD and improve quality of life.

Despite the promising evidence supporting lifestyle modifications, several barriers remain to their widespread adoption in clinical practice. Cancer survivors may face numerous challenges in implementing dietary and physical activity changes, including fatigue, psychological distress, and physical limitations. Therefore, healthcare providers should focus on individualized counseling and support to empower survivors to make sustainable changes. Furthermore, ongoing research is needed to identify the most effective strategies for promoting long-term adherence to lifestyle interventions in cancer survivors.

## 5. Conclusion

In conclusion, the increasing population of cancer survivors presents an urgent need for comprehensive cardiovascular care to address the long-term risks associated with cancer therapies. The evidence from our meta-analysis supports the growing recognition of cancer therapy-induced cardiotoxicity as a significant contributor to heart failure and other cardiovascular conditions. While current guidelines provide some framework for cardiovascular monitoring, there is a clear need for more consistent implementation of these practices, as well as the development of evidence-based guidelines that incorporate a personalized approach to cardiovascular risk management. Lifestyle interventions, particularly dietary modifications and increased physical activity, offer a promising strategy to mitigate cardiovascular risks in this population. By integrating cardiovascular health monitoring into routine cancer survivorship care, we can improve the long-term health outcomes of cancer survivors and enhance their quality of life.

## Authors’ contributions

All authors contributed equally to the manuscript and read and approved the final version of the manuscript.

## Data Availability

Supplementary file

---

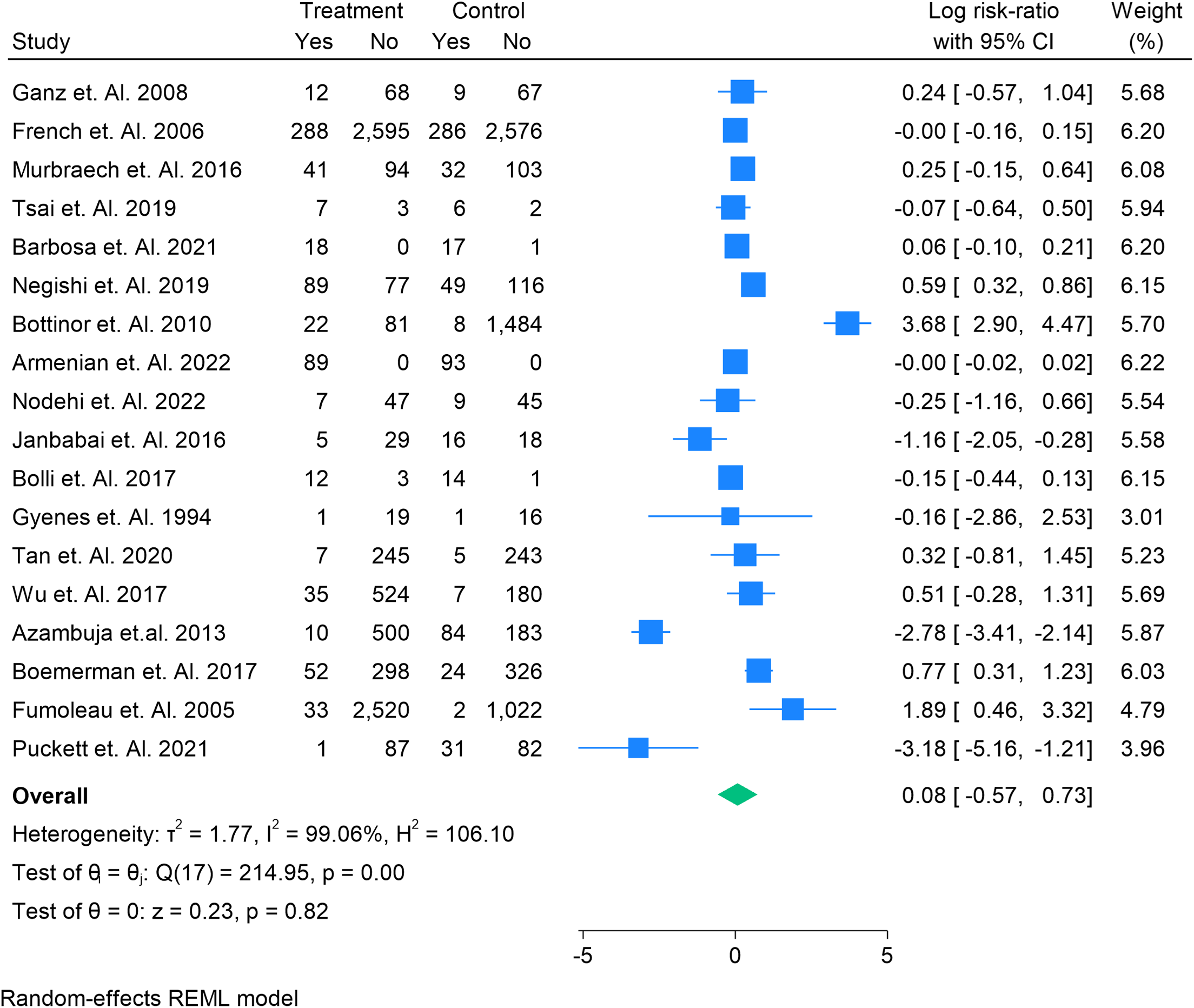

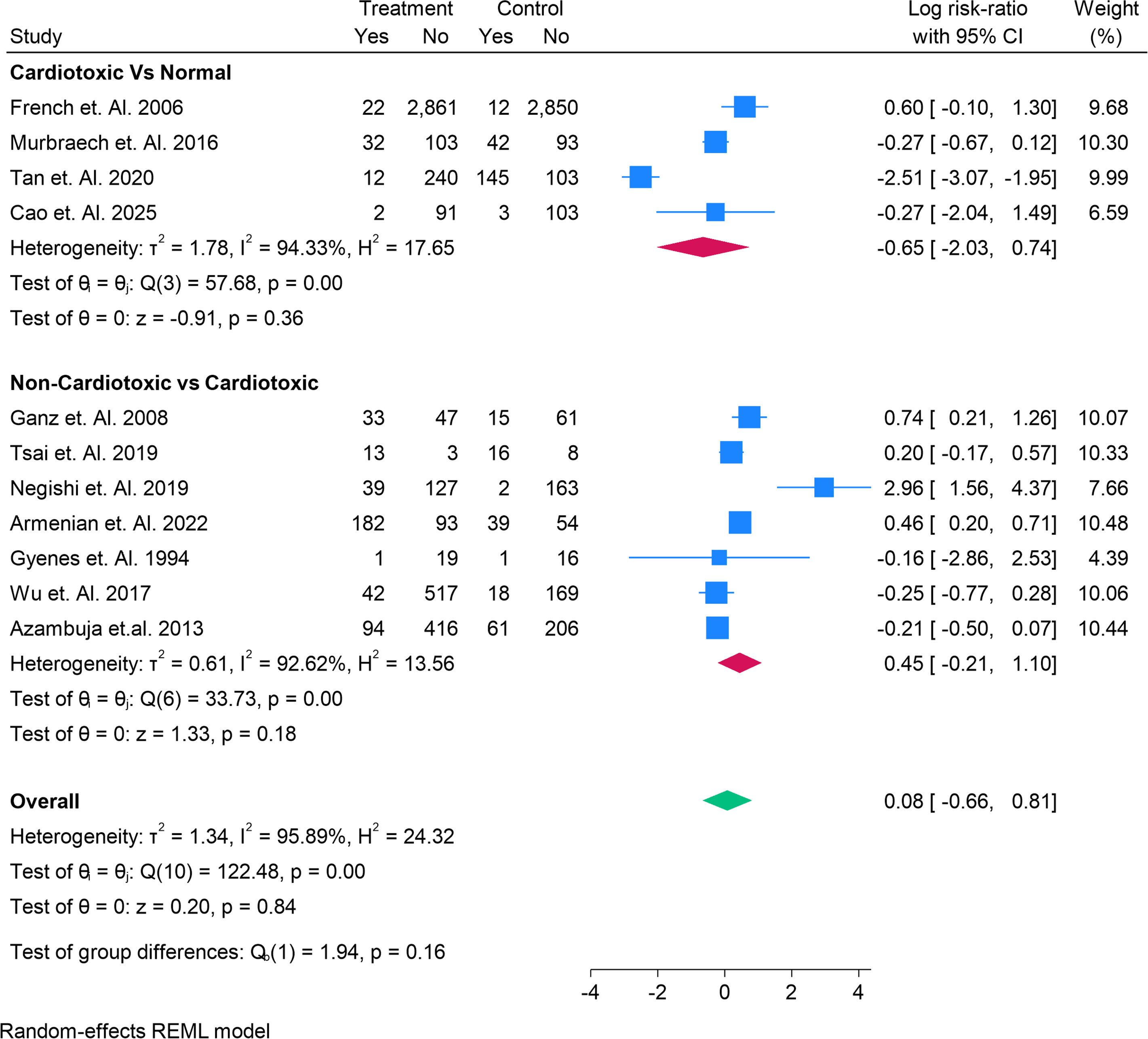

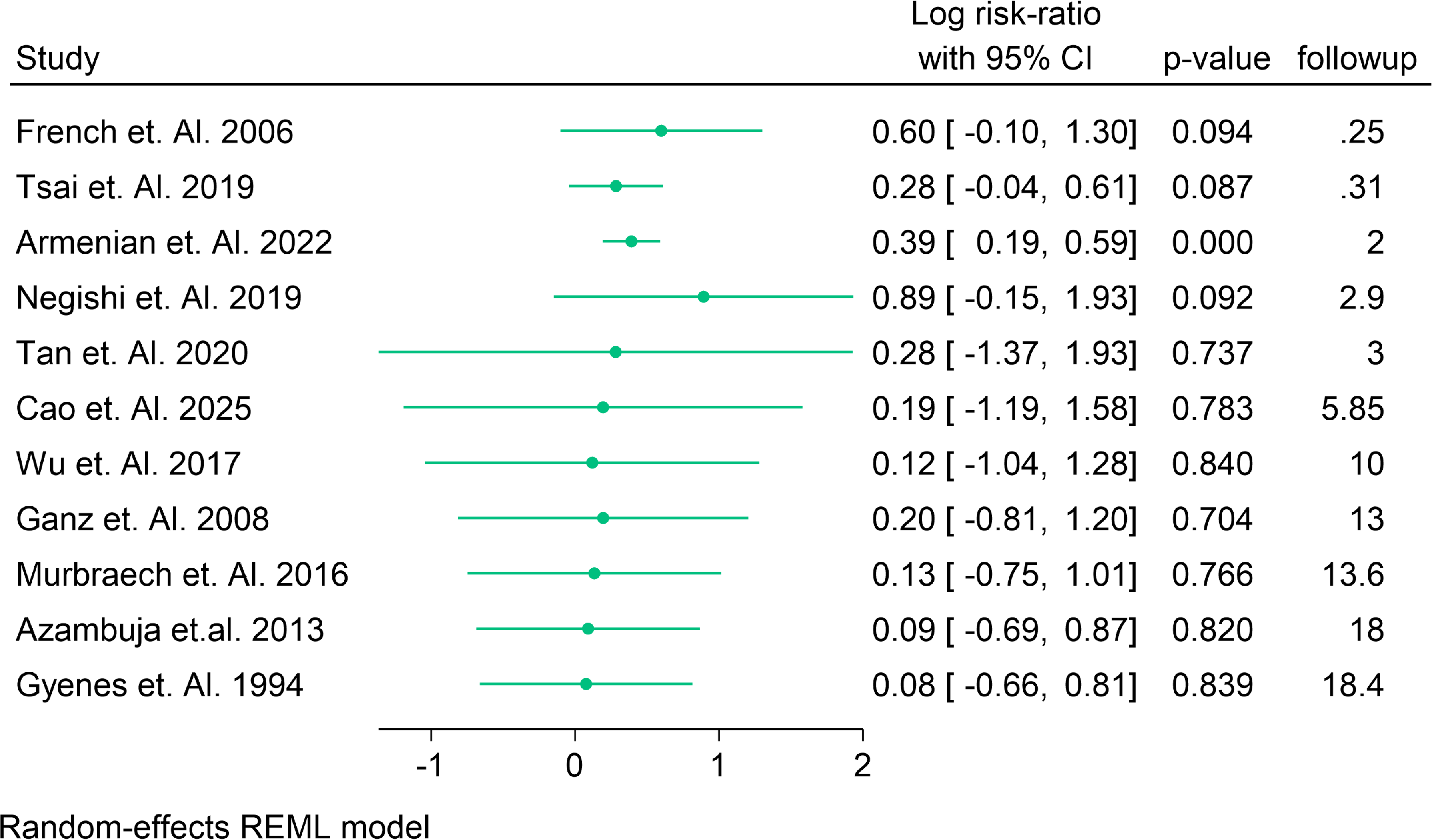

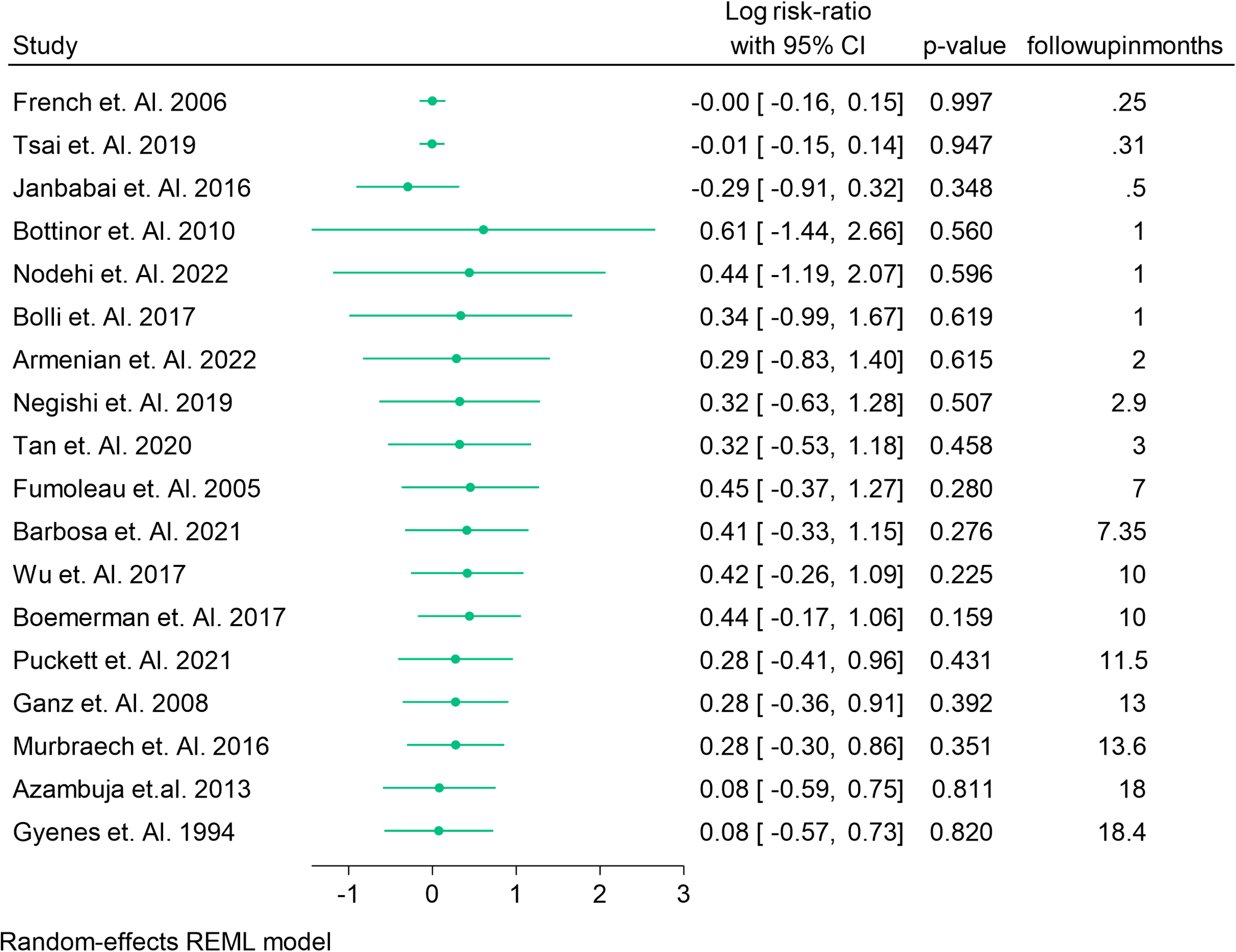

