## supplementary file for "Addressing the long-term cardiovascular risks of cardiotoxic therapies and emphasizing the critical role of lifestyle interventions in enhancing the health and well-being of cancer survivors: Systemic Review and Meta-analysis"

Supplementary Files

Search String

("heart failure" OR "cardiomyopathy" OR "congestive heart failure") AND ("cancer survivors" OR "post-cancer treatment") AND ("long-term outcomes" OR "late effects" OR "cardiotoxicity") AND ("chemotherapy" OR "radiation therapy" OR "targeted therapy" OR "immunotherapy") AND ("risk factors" OR "incidence" OR "prevalence") AND ("adult" OR "childhood cancer survivors")

Table S1. Details of All Studies

| **Ref Num** | **Author** | **Country** | **Population** | **Intervention** | **Control** | **Follow up** | **Average Age** | **Male Population** | **Female Population** | **GRADE** | **Main Finding** |
| --- | --- | --- | --- | --- | --- | --- | --- | --- | --- | --- | --- |
| 12 | Ganz et. Al. 2008 | USA | 180 | 80 | 76 | 13 | 55.6 | 0 | 180 | High | The study found no significant difference in the proportion of patients with reduced LVEF (less than 50%) between the CAF and CMF treatment groups at both 5-8 years and 10-13 years follow-up. The mean LVEF was slightly lower in the CAF group at 5-8 years, but not at 10-13 years. |
| 13 | French et. Al. 2006 | USA | 5745 | 2883 | 2862 | 0.25 | 60 | 3962 | 1783 | High | The study found that **mortality was high** in patients with **cardiogenic shock (CS)** and **congestive heart failure (CHF)** post-STEMI. Despite improvements in reperfusion therapy, these two conditions remained major causes of death, with poor outcomes through 90 days after the intervention. |
| 14 | Murbraech et. Al. 2016 | Norway | 270 | 135 | 135 | 13.6 | 56 | 170 | 104 | Moderate | The study found that lymphoma survivors treated with autologous hematopoietic stem cell transplantation (auto-HCT) had impaired right ventricular (RV) function, with more severe dysfunction in those who received high-dose cardiac radiotherapy. Left ventricular (LV) dysfunction was more prevalent, indicating a global cardiotoxic effect of the treatment. |
| 15 | Amioka et.al. 2013 | Japan | 249 | N/A | N/A | 2.8 | 67 | 121 | 128 | High | The major outcome was that **new-onset AF was associated with a higher incidence of acute heart failure and increased all-cause mortality**. The study concluded that **AF onset predicted unfavorable outcomes post-chemotherapy** |
| 16 | Tsai et. Al. 2019 | USA | 25 | 10 | 8 | 0.31 | 56.1 | 3 | 22 | High | Significant improvements in maximum oxygen uptake (V̇O 2max) and quality of life (QOL) functioning were observed in the intervention group. |
| 17 | Barbosa et. Al. 2021 | Brazil | 36 | 18 | 18 | 7.35 | 57.6 | 22 | 13 | High | In the treatment group, the Left Ventricular Ejection Fraction (LVEF) was slightly reduced (62.4 ± 7.5% compared to 68.0 ± 4.6% in the control group) |
| 18 | Negishi et. Al. 2019 | USA | 331 | 166 | 165 | 2.9 | 54 | 16 | 315 | High | The study found minimal difference in left ventricular ejection fraction (LVEF) between the EF-guided and GLS-guided groups over the 3-year period |
| 19 | Bottinor et. Al. 2010 | USA | 1572 | 103 | 1492 | 1 | 36 | 1100 | 471 | High | he study found that **adolescent and young adults (AYAs)** receiving **VEGF inhibitors** had a **lower incidence of hypertension** compared to non-AYAs. Additionally, the incidence of **LVSD** (left ventricular systolic dysfunction) was similar across both groups (AYAs and non-AYAs), independent of the therapy. |
| 20 | Armenian et. Al. 2022 | USA | 196 | 89 | 93 | 2 | 27.3 | 91 | 91 | High | The trial found that **low-dose carvedilol appeared to be safe** in childhood cancer survivors at high risk for heart failure due to anthracycline exposure, but it did not result in significant improvement in left ventricular wall thickness/dimension ratio (LVWT/Dz) compared to placeb |
| 21 | Bajestani et. Al. 2016 | Canada | 70 | N/A | N/A | 3 | 65.7 | 36 | 34 | High | The study highlighted that comprehensive cardiac evaluation in metastatic NSCLC patients revealed a high rate of undiagnosed cardiac abnormalities, including systolic heart failure, diastolic dysfunction, and other cardiovascular disorders, all of which could significantly affect the patients' performance status and cachexia status. This has implications for clinical trials assessing cachexia therapies in this population. |
| 22 | Nodehi et. Al. 2022 | Iran | 108 | 54 | 54 | 1 | 47.5 | 0 | 108 | Moderate | Non-inferiority of TA4415V compared to reference trastuzumab for efficacy in HER2-positive breast cancer patients in terms of pathologic complete response (pCR). The pCR rates were 37.5% in TA4415V and 34.09% in reference trastuzumab |
| 23 | Magnussen et. Al. 2017 | USA | 41830 | N/A | N/A | 10 | 26.3 | 32300 | 9530 | High | The study found that **survival after heart transplantation was comparable between women and men**, although there were **gender-specific differences** in graft failure, allograft vasculopathy, and malignancy. |
| 24 | Janbabai et. Al. 2016 | Iran | 69 | 34 | 34 | 0.5 | 47.4 | 5 | 64 | High | Enalapril significantly prevented the decrease in LVEF, preserving both systolic and diastolic function in cancer patients treated with anthracyclines. |
| 25 | Bolli et. Al. 2017 | USA | 36 | 15 | 15 | 1 | 48.5 | N/A | N/A | High | The primary outcomes are the **safety and feasibility** of allo-MSC therapy. Secondary outcomes include **changes in LV function** and **quality of life** measures. |
| 26 | Kim et. Al. 2021 | South Korea | 1256 | 628 | 628 | 4.1 | 51.4 | 0 | 1256 | High | The study validated a new cardiovascular risk score, the CHEMO-RADIAT score, which predicts major adverse cardiovascular events (MACE) after breast cancer treatment |
| 27 | Gyenes et. Al. 1994 | Sweden | 37 | 20 | 17 | 18.4 | 61.4 | N/A | N/A | High | The study found that **25% of the study group** (patients irradiated for left-sided disease) had **ischemic heart disease**, as identified by scintigraphy, whereas no cases were detected in the control group. |
| 28 | Gustavsson et. Al. 1999 | Sweden | 298 | 275 | 23 | 13 | 58 | 0 | 298 | High | **No serious cardiac sequelae** were observed after adjuvant radiotherapy following mastectomy in young women with early breast cancer, despite the use of older radiation techniques. |
| 29 | Tan et. Al. 2020 | USA | 500 | 252 | 248 | 3 | 49 | 0 | 500 | High | The trial's primary outcome was the **non-inferiority of subcutaneous pertuzumab** compared to intravenous pertuzumab for **HER2-positive early breast cancer** . |
| 30 | Wu et. Al. 2017 | USA | 746 | 559 | 187 | 10 | 48 | N/A | N/A | High | The **major outcome finding** was the increase in **arrhythmias** among patients treated with radiation therapy (RT) compared to those who did not receive RT, though there was no significant increase in the risk of **myocardial infarction** or **heart failure**. |
| 31 | Azambuja et.al. 2013 | Belgium | 777 | 510 | 267 | 18 | 47 | N/A | N/A | High | The study concluded that despite the higher doses of epirubicin in the treatment regimens (SDE/HDE), the incidence of long-term cardiac toxicity was relatively low in the cohort, suggesting that long-term monitoring may not be required for most patients. |
| 32 | Cao et. Al. 2025 | China | 199 | 93 | 106 | 5.85 | 52 | N/A | N/A | High | The study assessed the incidence of newly onset cardiac events within 1 year after radiotherapy, with no significant differences between the intervention and control groups . |
| 33 | Lee et. Al. 2017 | Taiwan | 3489 | 2376 | 1113 | 5.2 | 50.35 | N/A | 3489 | High | The study found that adjuvant radiotherapy (RT) and anthracycline-based chemotherapy (CT) can increase the risk of cardiotoxicity, especially when combined, and these therapies are significant independent prognostic factors for major heart events, particularly in younger patients with breast cancer. |
| 34 | Ryberg et. Al. 1998 | Denmark | 469 | 375 | 94 | 6.25 | 53 | N/A | N/A | High | The major outcome finding was the relationship between the cumulative dose of epirubicin and the risk of developing congestive heart failure (CHF). The risk increased exponentially from 4% at a cumulative dose of 900 mg/m² to 15% at 1,000 mg/m². Previous irradiation to the mediastinum and thoracic spine also increased the risk of CHF. |
| 35 | Shapiro et. Al. 1998 | USA | 552 | 276 | 276 | 19.4 | 50 | N/A | 575 | Moderate | The major finding was that **higher doses of adjuvant doxorubicin (CA10) were associated with a significantly increased risk of cardiac events**, particularly when combined with high dose-volume radiotherapy (RT). In contrast, **CA5** regimen did not significantly increase the risk of cardiac events compared to the general population. |
| 36 | Banke et. Al. 2019 | Denmark | 8701 | 2057 | 6644 | 5.4 | 52.34 | 0 | 8701 | High | Trastuzumab treatment after anthracycline-based chemotherapy is associated with a **two-fold increased risk of heart failure** (HF) compared to chemotherapy alone. The risk is particularly prominent in patients who had chemotherapy plus trastuzumab treatment after 18 months. |
| 37 | Boemerman et. Al. 2014 | Netherlands | 2196 | 561 | 1635 | 9 | 56 | 0 | 2196 | High | The study concluded that no significant increased risk of cardiovascular disease (CVD) was found among women treated for breast cancer with radiotherapy or chemotherapy, although the risk for congestive heart failure after chemotherapy (HR = 1.8) was in line with previous studies, but not statistically significant. |
| 38 | Boemerman et. Al. 2017 | Netherlands | 741 | 350 | 350 | 10 | 63 | 0 | 741 | High | Breast cancer survivors have a higher risk of mild left ventricular (LV) systolic dysfunction, elevated NT-proBNP levels, and cardiovascular disease (CVD) compared to matched controls |
| 39 | Chien et. Al. 2016 | Taiwan | 23006 | 1066 | 3794 | 5.9 | 50.99 | 0 | 23,006 | High | The primary outcome showed that trastuzumab use was associated with a significantly increased risk of heart failure and/or cardiomyopathy in the cohort, with a **hazard ratio (HR) of 1.86**. |
| 40 | Chung et. Al. 2020 | South Korea | 56,338 | 5643 | 6220 | 5.4 | 53.8 | 0 | 56338 | High | The study found that **standard low-dose anthracycline chemotherapy** significantly increases the risk of **late-onset congestive heart failure (CHF)** in breast cancer survivors, particularly those aged 50–59 years at diagnosis. |
| 41 | Franchi et. Al. 2020 | Italy | 28,599 | 6208 | 12416 | 5.88 | 55 | 0 | 28599 | High | The study finds that trastuzumab is associated with a higher short-term cardiovascular risk (especially heart failure and cardiomyopathy) during the treatment period. However, after cessation of the treatment, the long-term cardiovascular risk in patients treated with trastuzumab does not significantly differ from those treated with standard chemotherapy |
| 42 | Fumoleau et. Al. 2005 | France | 3577 | 2553 | 1024 | 7 | 59 | 0 |  | High | The study concluded that **epirubicin-based chemotherapy** had a **low risk** for long-term **left ventricular dysfunction** (LVD) and **heart failure**, with the risk being higher in older women and those with a higher BMI. |
| 43 | Ganz et. Al. 2017 | USA | 2119 | 1061 | 1058 | 8.8 | 50 | 0 | 2119 | High | The study found that the addition of trastuzumab to anthracycline-based chemotherapy did not result in long-term worsening of cardiac function, cardiac symptoms, or health-related quality of life in disease-free survivors. |
| 44 | Kwan et. Al. 2022 | USA | 88,838 | 14804 | 74034 | 7 | 61.1 | 0 | 88,838 | Moderate | Women with breast cancer have a 1.3-fold increased risk of developing heart failure (HF), both preserved (HFpEF) and reduced ejection fraction (HFrEF), compared to women without breast cancer. |
| 45 | Puckett et. Al. 2021 | USA | 201 | 88 | 113 | 11.5 | 50 |  |  | High | The major outcome of the study was that over half of the long-term breast cancer survivors showed evidence of clinical cardiovascular disease (CVD), with 77.6% exhibiting preclinical or clinical CVD. The study highlighted the importance of multi-modality screening for detecting CVD in these patients. |
| 46 | Ocier et. Al. 2021 | USA | 2129 |  |  | 5 | 65 | 1162 | 967 | High | Younger B-NHL survivors had higher relative risks of certain cardiovascular diseases, such as chronic rheumatic heart disease, myocarditis, diseases of the arteries, and hypotension, compared to older survivors and the general population. |

Table S2. Co-Factors Affecting the Survival in numbers in each co-factor

| Study Name | **Hypertension** | **Diabetes** | **Dyslipidemia** | **CKD** | **AF** | **Arrhythmia** | **IHD** | **AMI** | **HF** | **Smoking** | **COPD** | **CVD** | **ACEi** | **Beta-Blockers** | **Anti-Platelets** | **Statin** | **Diuretics** | **Surgery** | **Radiation** | **Left Breast** | **ChemoTherapy** | **Anthracycline** | **Hormone Therapy** | **Her2 Neu Inhibitors** | **Left Breast Surgery** | **Right Breast Surgery** |
| --- | --- | --- | --- | --- | --- | --- | --- | --- | --- | --- | --- | --- | --- | --- | --- | --- | --- | --- | --- | --- | --- | --- | --- | --- | --- | --- |
| Ganz et. Al. 2008 | 25 | 7 | 30 | N/A | N/A | 11 | N/A | N/A | N/A | 25 | N/A | N/A | N/A | N/A | N/A | N/A | N/A | N/A | 9 | N/A | 180 | 180 | 30 | N/A | N/A | N/A |
| French et. Al. 2006 | 3390 | 1320 | 3390 | 345 | 229 | 173 | 1381 | 4140 | 1090 | 2530 | 574 | 518 | 3731 | 5219 | 5115 | 3222 | 920 | 1494 | 57 | N/A | 288 | N/A | N/A | N/A | N/A | N/A |
| Murbraech et. Al. 2016 | 96 | 27 | 112 | N/A | N/A | N | N/A | N/A | 29 | 49 | N/A | N/A | 30 | 22 | N/A | 38 | N/A | N/A | 35 | N/A | 270 | 256 | N/A | N/A | N/A | N/A |
| Amioka et.al. 2013 | 74 | 33 | 32 | 7 | 15 | N | N/A | N/A | 15 | N/A | N/A | N/A | N/A | N/A | N/A | N/A | N/A | N/A | N | N/A | 249 | N/A | N/A | N/A | N/A | N/A |
| Tsai et. Al. 2019 | N/A | N/A | N/A | N/A | N/A | N/A | N/A | N/A | N/A | N/A | N/A | N/A | N/A | N/A | N/A | N/A | N/A | N/A | N | N/A | N/A | N/A | N/A | N/A | N/A | N/A |
| Barbosa et. Al. 2021 | 9 | 3 | 1 | N/A | N/A | N/A | N/A | N/A | N/A | N/A | N/A | N/A | N/A | N/A | N/A | N/A | N/A | N/A | N | N/A | N/A | N/A | N/A | N/A | N/A | N/A |
| Negishi et. Al. 2019 | 86 | 43 | 76 | N/A | N/A | N/A | N/A | N/A | N/A | 93 | N/A | N/A | 50 | 15 | N/A | N/A | N/A | N/A | 331 | N/A | 278 | N/A | N/A | N/A | N/A | N/A |
| Bottinor et. Al. 2010 | 811 | N/A | N/A | N/A | N/A | N/A | N/A | N/A | N/A | N/A | N/A | N/A | 238 | 141 | N/A | 15 | N/A | N/A | N/A | N/A | N/A | N/A | N/A | N/A | N/A | N/A |
| Armenian et. Al. 2022 | 8 | 8 | N/A | N/A | N/A | N | N/A | N/A | N/A | N/A | N/A | N/A | N/A | N/A | N/A | N/A | N/A | N/A | 34 | N/A | N/A | 196 | N/A | N/A | N/A | N/A |
| Bajestani et. Al. 2016 | 28 | 10 | N/A | N/A | N/A | N | N/A | N/A | N/A | N/A | N/A | N/A | N/A | N/A | N/A | N/A | N/A | N/A | N/A | N/A | N/A | N/A | N/A | N/A | N/A | N/A |
| Nodehi et. Al. 2022 | N/A | N/A | N/A | N/A | N/A | N | N/A | N/A | N/A | N/A | N/A | N/A | N/A | N/A | N/A | N/A | N/A | 18 | N/A | N/A | N/A | N/A | N/A | N/A | 42 | N/A |
| Magnussen et. Al. 2017 | N/A | N/A | N/A | N/A | N/A | N | N/A | N/A | N/A | N/A | N/A | N/A | N/A | N/A | N/A | N/A | N/A | N/A | N | N/A | N/A | N/A | N/A | N/A | N/A | N/A |
| Janbabai et. Al. 2016 | 10 | 8 | 7 | N/A | N/A | N | N/A | N/A | N/A | N/A | N/A | N/A | 69 | N/A | N/A | N/A | N/A | N/A | N | N/A | N/A | N/A | N/A | N/A | N/A | N/A |
| Bolli et. Al. 2017 | N/A | N/A | N/A | N/A | N/A | n | N/A | N/A | N/A | N/A | N/A | N/A | N/A | N/A | N/A | N/A | N/A | N/A | N | N/A | N/A | N/A | N/A | N/A | N/A | N/A |
| Kim et. Al. 2021 | 887 | N/A | N/A | N/A | N/A | N | N/A | N/A | N/A | N/A | N/A | N/A | N/A | N/A | N/A | N/A | N/A | N/A | 887 | 887 | 887 | N/A | 887 | 178 | N/A | N/A |
| Gyenes et. Al. 1994 | 13 | 7 | 20 | N/A | N/A | N | 5 | N/A | N/A | 13 | N/A | N/A | N/A | N/A | N/A | N/A | N/A | N/A | N | N/A | N/A | N/A | N/A | N/A | N/A | N/A |
| Gustavsson et. Al. 1999 | 9 | 1 | N/A | N/A | N/A | n | N/A | N/A | N/A | N/A | N/A | N/A | N/A | N/A | N/A | N/A | N/A | N/A | n | 139 | 298 | N/A | N/A | N/A | 139 | 136 |
| Tan et. Al. 2020 | N/A | N/A | N/A | N/A | N/A | n | N/A | N/A | N/A | N/A | N/A | N/A | N/A | N/A | N/A | N/A | N/A | N/A | n | N/A | 500 | 495 | 382 | 500 | N/A | N/A |
| Wu et. Al. 2017 | 134 | 15 | 22 | 15 | N/A | 15 | 12 | 4 | 18 | 7 | N/A | N/A | N/A | N/A | N/A | N/A | N/A | N/A | N | N/A | 559 | N/A | N/A | N/A | N/A | N/A |
| Azambuja et.al. 2013 | 425 | 170 | N/A | N/A | N/A | N | N/A | N/A | N/A | N/A | N/A | N/A | N/A | N/A | N/A | N/A | N/A | N/A | N | N/A | N/A | N/A | N/A | N/A | N/A | N/A |
| Cao et. Al. 2025 | 35 | 6 | 7 | N/A | N/A | N/A | N/A | N/A | N/A | N/A | N/A | N/A | N/A | N/A | N/A | N/A | N/A | N/A | 199 | 119 | 199 | 143 | N/A | 64 | 119 | 80 |
| Lee et. Al. 2017 | 720 | 238 | N/A | N/A | N/A | N | N/A | N/A | 303 | N/A | N/A | N/A | N/A | N/A | N/A | N/A | N/A | N/A | 1671 | N/A | 1730 | N/A | N/A | N/A | N/A | N/A |
| Ryberg et. Al. 1998 | N/A | N/A | N/A | N/A | N/A | N | N/A | N/A | 34 | N/A | N/A | N/A | N/A | N/A | N/A | N/A | N/A | N/A | 176 | 92 | N/A | 469 | N/A | N/A | N/A | N/A |
| Shapiro et. Al. 1998 | N/A | N/A | N/A | N/A | N/A | N | N/A | N/A | 19 | N/A | N/A | N/A | N/A | N/A | N/A | N/A | N/A | 507 | 122 | 65 | 552 | 552 | N/A | N/A | 65 | 50 |
| Banke et. Al. 2019 | 663 | 144 | N/A | N/A | 44 | N | 178 | 33 | N/A | N/A | 94 | N/A | N/A | N/A | N/A | N/A | N/A | 4145 | 5315 | N/A | 343.4 | N/A | N/A | N/A | N/A | N/A |
| Boemerman et. Al. 2014 | 263 | 72 | 106 | N/A | N/A | N | 30 | N/A | N/A | N/A | N/A | N/A | N/A | N/A | N/A | N/A | N/A | 561 | 229 | 107 | 145 | N/A | 49 | 4 | 107 | 120 |
| Boemerman et. Al. 2017 | N/A | N/A | 54 | N/A | 11 | N | 26 | N/A | 4 | N/A | N/A | N/A | 65 | 54 | 29 | 54 | 33 | N/A | 240 | 284 | 284 | 218 | N/A | 26 | N/A | N/A |
| Chien et. Al. 2016 | 4503 | 2068 | 641 | 300 | N/A | N | N/A | N/A | N/A | N/A | N/A | N/A | 1250 | 4880 | 2070 | 1920 | 4380 | N/A | 254 | 1145 | N/A | N/A | N/A | N/A | N/A | N/A |
| Chung et. Al. 2020 | 3045 | 846 | 3687 | N/A | N/A | N | N/A | N/A | 124 | N/A | N/A | N/A | N/A | N/A | N/A | N/A | N/A | N/A | 8356 | N/A | N/A | N/A | N/A | N/A | N/A | N/A |
| Franchi et. Al. 2020 | 8453 | 2344 | N/A | 57 | N/A | 200 | N/A | N/A | N/A | N/A | N/A | N/A | N/A | N/A | N/A | N/A | N/A | N/A | 21,499 | N/A | N/A | N/A | N/A | N/A | N/A | N/A |
| Fumoleau et. Al. 2005 | N/A | N/A | N/A | N/A | N/A | n | N/A | N/A | 5 | N/A | N/A | N/A | N/A | N/A | N/A | N/A | N/A | N/A | N/A | N/A | N/A | N/A | N/A | N/A | N/A | N/A |
| Ganz et. Al. 2017 | 451 | 81 | 188 | N/A | 68 | 118 | N/A | 17 | 19 | 797 | N/A | N/A | N/A | N/A | N/A | N/A | N/A | N/A | 1303 | N/A | N/A | N/A | N/A | N/A | N/A | N/A |
| Kwan et. Al. 2022 | 6240 | 1997 | 6434 | 3778 | N/A | 143 | 4837 | N/A | N/A | N/A | N/A | N/A | N/A | N/A | N/A | N/A | N/A | N/A | 4813 | N/A | 6229 | 4112 | 7874 | N/A | N/A | N/A |
| Puckett et. Al. 2021 | 53 | 30 | 72 | 1 | 5 | 5 | N/A | 104 | 61 | N/A | N/A | 3 | N/A | N/A | N/A | N/A | N/A | N/A | n | N/A | N/A | N/A | N/A | N/A | N/A | N/A |
| Ocier et. Al. 2021 | N/A | N/A | N/A | N/A | N/A | n | N/A | N/A | N/A | 128 | N/A | N/A | N/A | N/A | N/A | N/A | N/A | N/A | n | N/A | N/A | N/A | N/A | N/A | N/A | N/A |
